# Post-stroke changes in the ipsilesional motor area after contralesional continuous theta-burst stimulation

**DOI:** 10.1101/2024.11.20.24317674

**Authors:** Jord JT Vink, Camille FM Biemans, Eline CC van Lieshout, Ruben PA van Eijk, Sebastiaan FW Neggers, Johanna MA Visser-Meily, H Bart van der Worp, Rick M Dijkhuizen

## Abstract

**Background:** Inhibitory repetitive transcranial magnetic stimulation (rTMS) of the contralesional primary motor cortex (M1) can promote upper limb recovery after stroke. However, its working mechanism remains unclear. We hypothesised that contralesional inhibitory rTMS increases ipsilesional M1 excitability and promotes ipsilesional motor-eloquent area (MEA) remapping.

**Methods:** Sixty patients who participated in a trial on contralesional continuous theta-burst stimulation (cTBS), an inhibitory form of rTMS, for the promotion of upper limb recovery after stroke, were included. M1 excitability and upper limb function were measured from TMS-based resting motor thresholds (RMTs) and the Fugl-Meyer Assessment (FMA) arm score, respectively, before cTBS treatment and at six follow-up visits up to one year after stroke. Forty-four patients additionally underwent longitudinal navigated TMS-based motor mapping. Remapping of the MEA was assessed from longitudinal changes in MEA overlap. Outcomes were analysed using mixed models for repeated measures.

**Results:** The ipsilesional RMT was 11% lower after active cTBS compared to sham cTBS (95% CI −18.7 to −2.6; p 0.0099) within twelve hours after the series of treatments. Compared to the sham cTBS group, MEA overlap was 27% (95% CI −44 to −11; p 0.0030), 25% (95% CI −45 to −5; p 0.0224) and 29% (95% CI −48 to −11; p 0.0038) less in the active cTBS group within twelve hours and at one week post-treatment, and three months post-stroke, respectively. Ipsilesional M1 excitability (i.e., RMT) within twelve hours post-cTBS correlated with FMA arm score at 3 months post-stroke (Spearman’s rho 0.59; p < 0.0001).

**Conclusions:** Upper limb recovery after cTBS treatment of the contralesional M1 after stroke may be caused by increased ipsilesional M1 excitability and MEA remapping.

**Trial registration:** https://trialsearch.who.int/; Unique identifier: NTR6133.

## Introduction

It has become increasingly apparent that recovery of motor function during rehabilitation after stroke is associated with plasticity of the motor cortex.^1^ Physical therapy as applied during stroke rehabilitation may promote functional remapping of the primary motor cortex (M1) and has been shown to lead to the expansion of the motor-eloquent cortex into neighbouring cortex in animal models.^2,3^ However, the underlying neurophysiological mechanisms of therapy-induced functional recovery in patients after stroke remain poorly understood.^4–7^

Transcranial magnetic stimulation (TMS) is a non-invasive method that enables assessment of the neurophysiological underpinnings of post-stroke upper limb recovery.^8,9^ TMS of the motor cortex can evoke a muscular response, which can be measured as a motor-evoked potential (MEP) with electromyography (EMG).^9^ Single-pulse TMS in combination with EMG and MRI-guided neuronavigation allows localisation of the motor-eloquent area (MEA).^10^ Applied longitudinally, this approach can be used to detect changes in the topological representation of the MEA to identify MEA remapping.^11^ TMS-based motor mapping has previously revealed an increase in MEA size after stroke, which was associated with spontaneous or constraint-induced movement therapy (CIMT)-induced recovery of the upper limb.^9,12^ A MEA shift, on the other hand, is variable and a relationship with upper limb recovery has not been established.^9^

When applied repetitively, TMS may up or downregulate cortical excitability in the targeted area, depending on the stimulation paradigm. Inhibitory repetitive TMS (rTMS) of the contralesional M1 combined with physical therapy, started within the first month after stroke onset, can improve upper limb recovery in patients.^13–15^ Inhibitory rTMS can be performed with low-frequency (LF) rTMS or continuous theta-burst stimulation (cTBS).^16^ Upper limb recovery after stroke has been associated with an increase in ipsilesional M1 excitability, reducing or eliminating the interhemispheric imbalance (i.e. asymmetry) in M1 excitability.^16^ Therefore, a potential mode of action of LF rTMS or cTBS of the contralesional M1 is restoration of the balance of interhemispheric interactions, through downregulation of contralesional M1 excitability and potentially (consequent) upregulation of ipsilesional M1 excitability.^17,18^ However, the relationship between (rTMS-induced) restoration of the interhemispheric balance in M1 excitability, MEA remapping and upper limb recovery remains unclear.

In a randomised trial including 60 patients, we previously showed that ten sessions of cTBS of the contralesional M1, combined with physical therapy of the affected arm, delivered within the first three weeks after stroke promotes upper limb recovery.^15^ Excitability measurements showed that cTBS may have a suppressive effect on contralesional M1 excitability.^15^ We therefore hypothesise that ten sessions of contralesional cTBS led to an increase in ipsilesional M1 excitability, as defined by a larger reduction in the ipsilesional resting motor threshold (RMT) compared to sham cTBS after treatment, in the same patient cohort. Furthermore, we hypothesise that contralesional cTBS resulted in remapping of the ipsilesional MEA, as defined by a larger reduction in overlap with the baseline MEA compared to sham cTBS. Finally, we hypothesise that MEA remapping, as defined by a longitudinal reduction in overlap with the baseline MEA, was restricted to the ipsilesional MEA.

## Methods

### Study design

This study was part of a randomised, sham-controlled clinical trial on early cTBS treatment over the contralesional M1 for the promotion of upper limb recovery after stroke (B-STARS), B-STARS was performed according to the CONSORT (Consolidated Standards of Reporting Trials) guidelines and was approved by the Medical Research Ethics Committee of the University Medical Centre Utrecht.^15^ A summary of the trial protocol has been published^19^ and the full protocol is available in the supplemental material. The current study analyses secondary outcome measures of the B-STARS trial and relationships between neurophysiological and functional outcomes. Deidentified participant data are available from the corresponding author upon reasonable request.

### Participants

Patients were recruited from rehabilitation centre De Hoogstraat (Utrecht, the Netherlands). Patients who met the following inclusion criteria were eligible for participation: (1) age ≥18 years; (2) first-ever ischaemic stroke or intracerebral haemorrhage; (3) paresis of one arm, as determined by a Motricity Index (MI) of the arm between 9 and 99; (4) baseline assessment within three weeks after stroke onset. Patients were excluded from participation based on the following criteria: (1) disabling medical history as determined by the treating physician; (2) normal to almost normal use of the hand; (2) severe deficits in communication, memory, or understanding that impede proper study participation; and (3) contraindications to TMS as based on TMS safety guidelines.^20^ Acquisition of motor mapping data started after the 16^th^ patient was included in the B-STARS trial.

### Randomisation and masking

Patients were randomly assigned to ten daily sessions of contralesional cTBS or sham cTBS applied during two weeks in addition to regular care upper limb therapy, using a secured online allocation system (Research Online V2.0, Julius Centre, the Netherlands). Randomisation was stratified according to the ability to extend one or more fingers of the paretic arm.^21^ Sham cTBS was performed at 10% of the RMT (see below), with the TMS coil rotated 45 degrees relative to the scalp, and patients were masked to treatment allocation using auditory masking of the TMS coil sound.

### Procedures

#### TMS intervention

Treatment was delivered in ten daily sessions over the ‘motor hotspot’ of the contralesional M1, started within three weeks after stroke onset. The motor hotspot was defined as the position on the scalp at which MEPs with the largest peak-to-peak amplitude could be evoked in the contralateral first dorsal interosseous (FDI) muscle by delivering TMS pulses and monitoring the electromyogram. The RMT was determined before each treatment. The RMT was defined as the minimum machine output (MO) at which stimulation evoked at least five out of ten MEPs with a peak-to-peak amplitude of over 50 μV.^22^ The RMT was identified by increasing the MO in steps of 5 and 10% for the contralesional and ipsilesional M1, respectively, until an MEP was detected, and reduced with steps of 1% until the RMT was determined. A Neuro-MS/D advanced therapeutic magnetic stimulator and an angulated 100-mm figure-of-eight TMS coil (Neurosoft, Ivanovo, Russia) were used for stimulation. EMG was recorded, amplified, and digitised at a sampling frequency of 20 kHz using a four-channel Neuro-MEP amplifier (Neurosoft, Ivanovo, Russia). cTBS consisted of continuous delivery of three stimuli bursts at 50 Hz repeated at five bursts per second for a duration of forty seconds with a biphasic TMS-induced current at 45 degrees to the midline. Stimulation intensity was set at 70% of the RMT.

cTBS or sham cTBS was delivered 15 minutes before daily standard upper limb therapy, which consisted of a 60-minute group therapy session of individualised upper limb exercises, according to the Concise Arm and hand Rehabilitation Approach in Stroke (CARAS).^23^ CARAS consists of a daily training programme during which patients perform specific exercises with the paretic arm under guidance of physical or occupational therapists, supplemented with exercises they can perform independently during the rest of the day.

#### TMS motor mapping

Navigated TMS (nTMS) motor mapping was performed at baseline (within three weeks after stroke), at the final (10^th^) day of the treatment period, at one week and one month after treatment, and at three, six and twelve months after stroke onset.

The motor hotspot and RMT were identified according to previously described procedures, using the same equipment. The motor hotspot was identified for each session individually, as the motor hotspot may have shifted due to neuroplastic changes.

Neuronavigation was performed on a generic MRI head model using the Neural Navigator neuronavigation system (Brain Science Tools, De Bilt, the Netherlands). Accurate registration of the MRI head model with the patient’s head across sessions was ensured by adjusting the alignment markers in the MRI head model based on the position of the facial landmarks captured during the alignment procedure. Registration errors of subsequent sessions were kept below 4 mm. This approach allowed registration of the MRI head model with a post-hoc acquired individual MRI scan. This was performed by refitting the facial landmarks measured before treatment, with the same locations marked in the post-hoc acquired individual MRI scan. A predefined grid of points was placed over the scalp, spaced 8 mm apart in both directions, covering the entire scalp. The neuronavigation system was used to guide the TMS coil to the predefined grid points.

During the baseline session, a motor map was acquired by delivering five TMS pulses at 120%RMT to each point of a 4-by-4 grid centered around the motor hotspot, totaling 80 stimuli. During subsequent sessions, the same number of TMS pulses at the same intensity were delivered to points of the same 4-by-4 grid (figure 1). This was done to ensure that the location of the grid would not explain longitudinal MEA changes.

**Figure 1.**
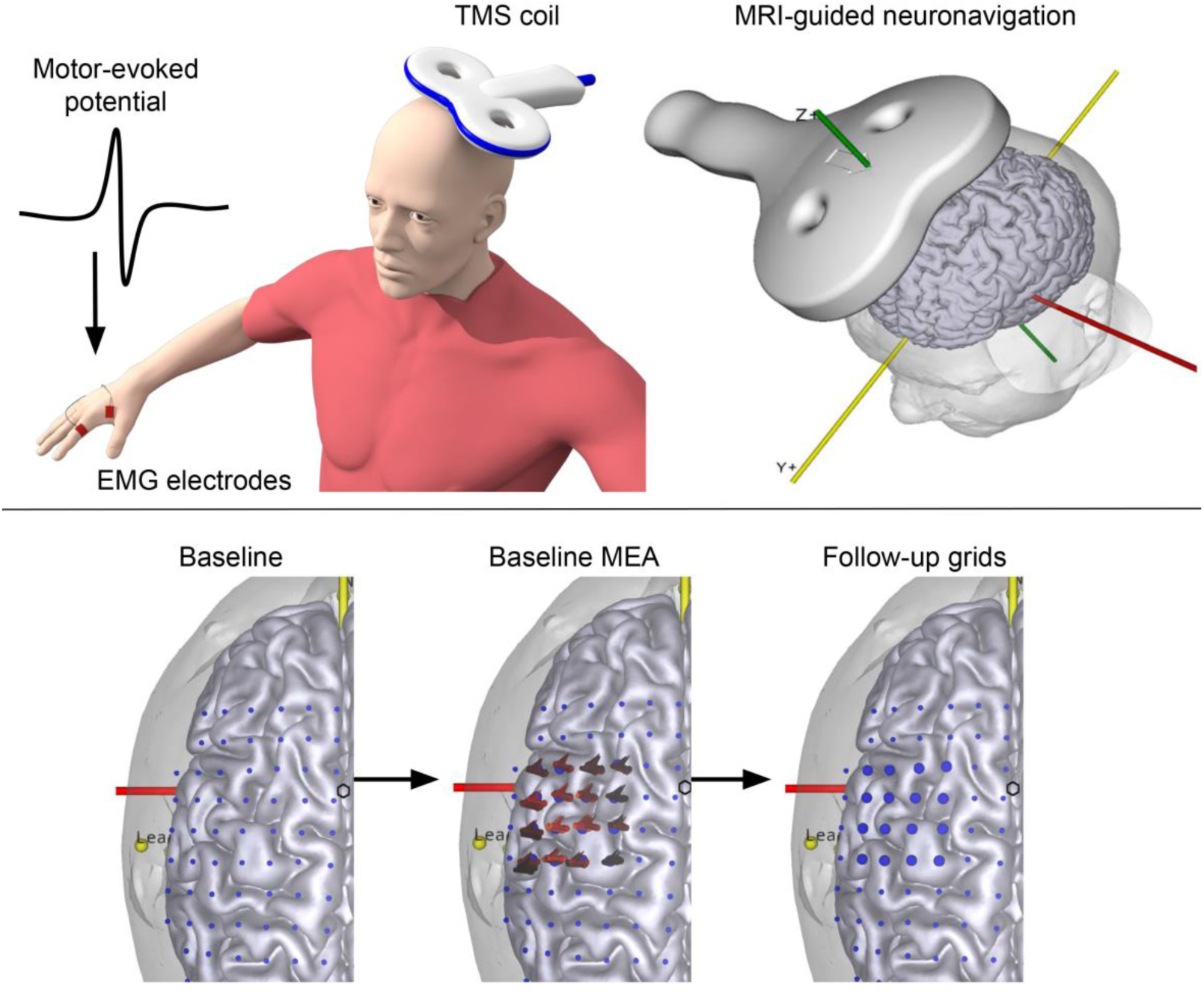
Schematic overview of the motor mapping procedure. Motor potentials evoked by TMS pulses delivered over M1 were recorded in the FDI muscle for both hemispheres. TMS coil placement was guided and recorded using an MRI-guided neuronavigation system. At baseline, the motor-eloquent area (MEA) was sampled in a 4-by-4 grid around the motor hotspot. The same grid points were stimulated at follow-up visits. The locations of the flags indicate the intersection of the TMS coil isocentre with the cortex, while the direction of the flag indicates the direction of the TMS-induced current. A brighter flag colour indicates a higher MEP amplitude evoked in the FDI muscle.

#### Upper limb function assessment

Upper limb function was assessed from the Fugl-Meyer Assessment (FMA) arm score at baseline (within three weeks after stroke), at one week and one month after treatment, and at three, six and twelve months after stroke onset by trained assessors.

The FMA arm score is a reliable and valid motor performance test consisting of 33 tasks that involve movements of the affected upper limb, with higher scores indicating better performance.^17^ Upper limb recovery was measured from the change in FMA arm score between the baseline and follow-up visits.

#### Data analysis

Data were analysed using custom scripts in MATLAB environment (MathWorks Inc., United States). Cortical excitability was defined by the RMT value. MEA remapping was defined from the change in overlap between the first measured MEA (depending on the presence of MEPs) and subsequent measured MEAs. In healthy individuals the overlap in MEAs has been shown to have a higher test-retest reliability as compared to centre of gravity (COG) shifts, making it more reliable for measurement of longitudinal changes in MEA representation.^11^ Percent overlap was calculated using the following formula:^24^

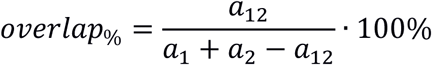

*a*_12_: the number of common grid points at which an average MEP with amplitude of >50 µV could be identified across two visits; and *a*_1_ and *a*_2_: the number of grid points at which an average MEP with amplitude of >50 µV could be measured during the first and second visit, respectively. An overlap of 100% indicates MEAs with identical spatial representation.

MEA remapping was additionally analysed from MEA size change and COG displacement over time. MEA size was calculated by multiplying the number of grid points at which an average MEP with an amplitude of more than 50 µV could be measured with the area covered by a single grid point (0.64 cm^2^). MEA displacement was measured from the displacement of the COG of the spatial distribution of MEP amplitudes in the MEA, along the mediolateral (ML) and anteroposterior (AP) axes, between the first visit (depending on the presence of MEPs) and subsequent visits. COGs were calculated by the following formulas:^25^

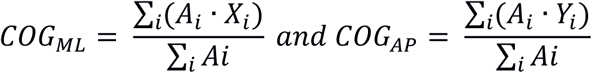

*Xi* and *Yi*: the coordinates of stimulation target *i* along the ML axis and the AP axis, respectively; *Ai*: amplitude of the MEP evoked at stimulation target *i*. COG displacement was assessed along the individual axes and as the Euclidian displacement.

#### Statistical analysis

The current analysis is an analysis of additional secondary outcomes. Statistical analysis of these additional secondary outcomes was performed according to the prespecified statistical analysis plan, which is available in the Supplemental Material and was completed before data-lock. Statistical analysis was performed using R 4.1. All hypotheses were tested two-tailed with an alpha of 0.05. Missing data were assumed to be missing at random. Analyses on M1 excitability were performed in all 60 randomised participants. The other analyses were performed on the 44 patients who underwent TMS-based motor mapping.

We investigated the effect of treatment on the ipsilesional RMT and MEA overlap at different visits. We investigated this using a mixed model for repeated measures (MMRM) with an unstructured variance-covariance matrix with visit (<12 hours, one week and one month post-treatment and three, six and twelve months post-stroke) and the interaction between visit and treatment (active and sham cTBS). The model with M1 excitability as outcome also included the baseline value. The effect of treatment on MEP status was analysed using a cumulative link mixed model. We performed a sensitivity analysis in which an additional covariate (i.e. baseline MEP status) was included, to assess potential bias introduced by MEP status.

Pearson’s correlation coefficients, or Spearman’s correlation coefficients in case of non-normally distributed data, were calculated to evaluate the relationship between the neurophysiological outcomes and upper limb recovery outcomes.

## Results

Between April 14, 2017 and February 12, 2021, a total of 494 stroke patients with arm weakness were screened for eligibility, of whom 60 were enrolled (Figure 2). Twenty-nine patients were randomly assigned to receive active cTBS, of whom one withdrew consent before starting treatment, resulting in 28 patients in the cTBS group, and 31 patients were assigned to receive sham cTBS. All 60 patients underwent cortical excitability and motor function measurements. The baseline characteristics and patient flow diagram of the full patient population have been reported previously.^15^ Forty-four out of sixty patients enrolled in the B-STARS study underwent additional motor mapping measurements. Of these forty-four patients, twenty-one patients were randomised to receive active cTBS, and twenty-three were assigned to receive sham cTBS. Not all patients could undergo motor mapping measurements at all visits due to temporary suspension of study activities as a result of the COVID-19 pandemic. A single patient showed signs of syncope during RMT determination and was excluded from the remaining motor mapping sessions. No serious adverse events (SAEs) were reported during the motor mapping sessions.

**Figure 2.**
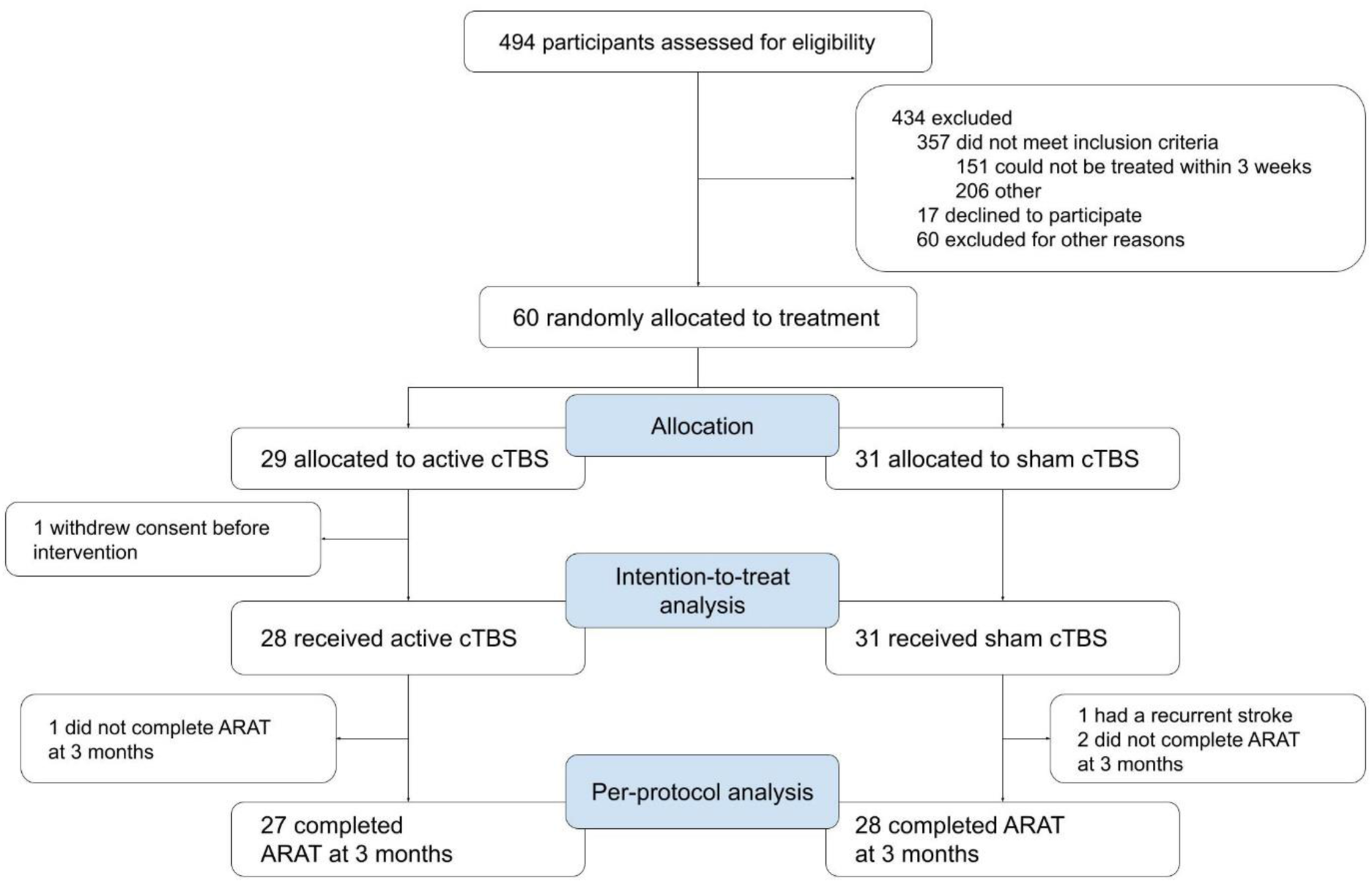
Participant flow diagram

**Table 1.**
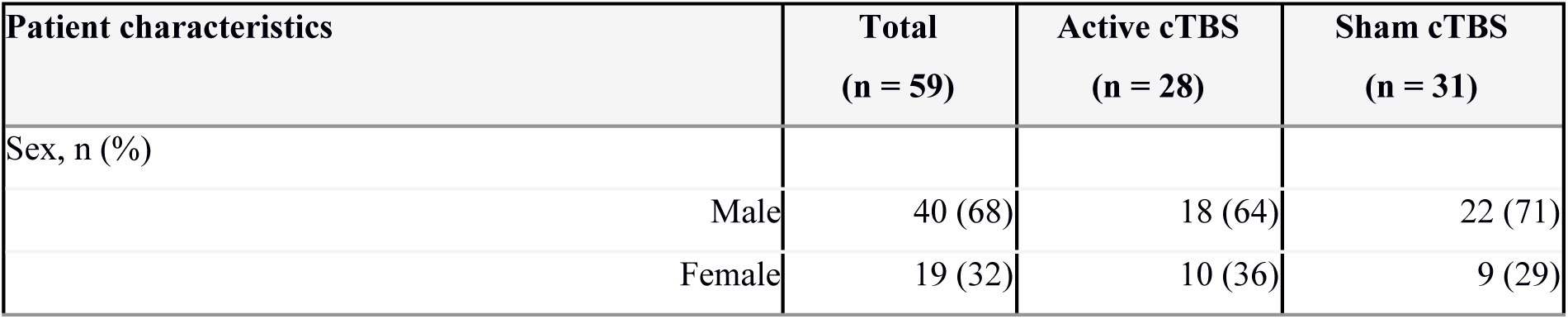
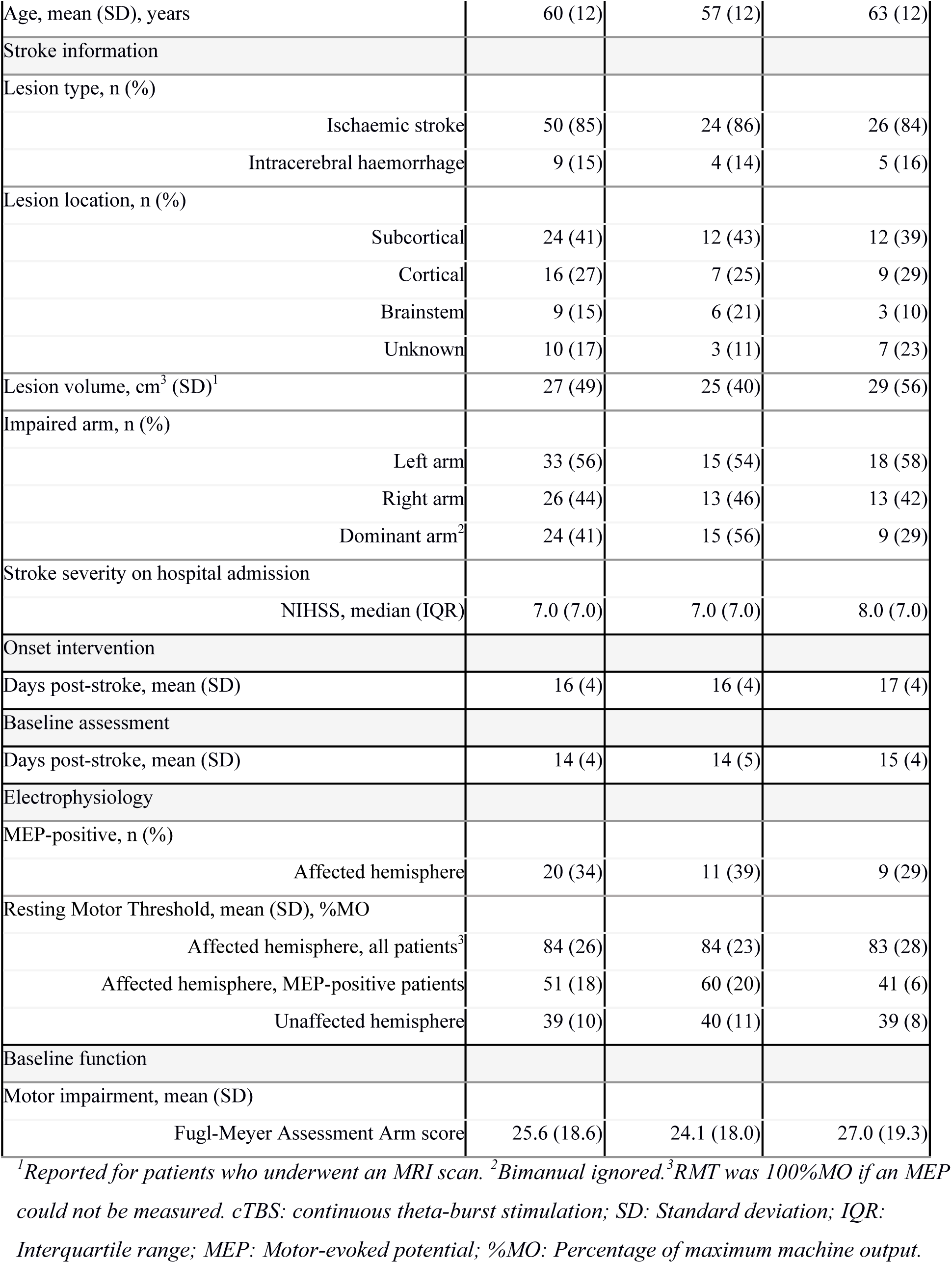
Baseline characteristics.

| Patient characteristics |  | Total<br>(n = 59) | Active cTBS<br>(n = 28) | Sham cTBS<br>(n = 31) |
| --- | --- | --- | --- | --- |
| Sex, n (%) |  |  |  |  |
|  | Male | 40 (68) | 18 (64) | 22 (71) |
|  | Female | 19 (32) | 10 (36) | 9 (29) |

|  |  |  |  |
| --- | --- | --- | --- |
| Age, mean (SD), years | 60 (12) | 57 (12) | 63 (12) |
| Stroke information |  |  |  |
| Lesion type, n (%) |  |  |  |
| Ischaemic stroke | 50 (85) | 24 (86) | 26 (84) |
| Intracerebral haemorrhage | 9 (15) | 4 (14) | 5 (16) |
| Lesion location, n (%) |  |  |  |
| Subcortical | 24 (41) | 12 (43) | 12 (39) |
| Cortical | 16 (27) | 7 (25) | 9 (29) |
| Brainstem | 9 (15) | 6 (21) | 3 (10) |
| Unknown | 10 (17) | 3 (11) | 7 (23) |
| Lesion volume, cm<sup>3</sup> (SD)<sup>1</sup> | 27 (49) | 25 (40) | 29 (56) |
| Impaired arm, n (%) |  |  |  |
| Left arm | 33 (56) | 15 (54) | 18 (58) |
| Right arm | 26 (44) | 13 (46) | 13 (42) |
| Dominant arm<sup>2</sup> | 24 (41) | 15 (56) | 9 (29) |
| Stroke severity on hospital admission |  |  |  |
| NIHSS, median (IQR) | 7.0 (7.0) | 7.0 (7.0) | 8.0 (7.0) |
| Onset intervention |  |  |  |
| Days post-stroke, mean (SD) | 16 (4) | 16 (4) | 17 (4) |
| Baseline assessment |  |  |  |
| Days post-stroke, mean (SD) | 14 (4) | 14 (5) | 15 (4) |
| Electrophysiology |  |  |  |
| MEP-positive, n (%) |  |  |  |
| Affected hemisphere | 20 (34) | 11 (39) | 9 (29) |
| Resting Motor Threshold, mean (SD), %MO |  |  |  |
| Affected hemisphere, all patients<sup>3</sup> | 84 (26) | 84 (23) | 83 (28) |
| Affected hemisphere, MEP-positive patients | 51 (18) | 60 (20) | 41 (6) |
| Unaffected hemisphere | 39 (10) | 40 (11) | 39 (8) |
| Baseline function |  |  |  |
| Motor impairment, mean (SD) |  |  |  |
| Fugl-Meyer Assessment Arm score | 25.6 (18.6) | 24.1 (18.0) | 27.0 (19.3) |

<sup>1</sup>Reported for patients who underwent an MRI scan. <sup>2</sup>Bimanual ignored. <sup>3</sup>RMT was 100%MO if an MEP could not be measured. cTBS: continuous theta-burst stimulation; SD: Standard deviation; IQR: Interquartile range; MEP: Motor-evoked potential; %MO: Percentage of maximum machine output.

The ipsilesional RMT was 10.6% lower in the active cTBS group compared to the sham cTBS group (95% CI −18.7 to −2.6; p 0.0099) within twelve hours after the series of treatments (Figure 3A). This difference was −8.2% at 1 week after treatment (95% CI −16.5 to 0.1; p 0.0533). After that, the ipsilesional RMT in both groups gradually converged over time. Sensitivity analysis including baseline MEP status showed that the ipsilesional RMT was 12.1% lower in the active group within twelve hours after treatment (95% CI −20.6 to −3.7; p 0.0050) and −9.6% at 1 week after treatment (95% CI −18.3 to −1.0; p 0.0296).

**Figure 3.**
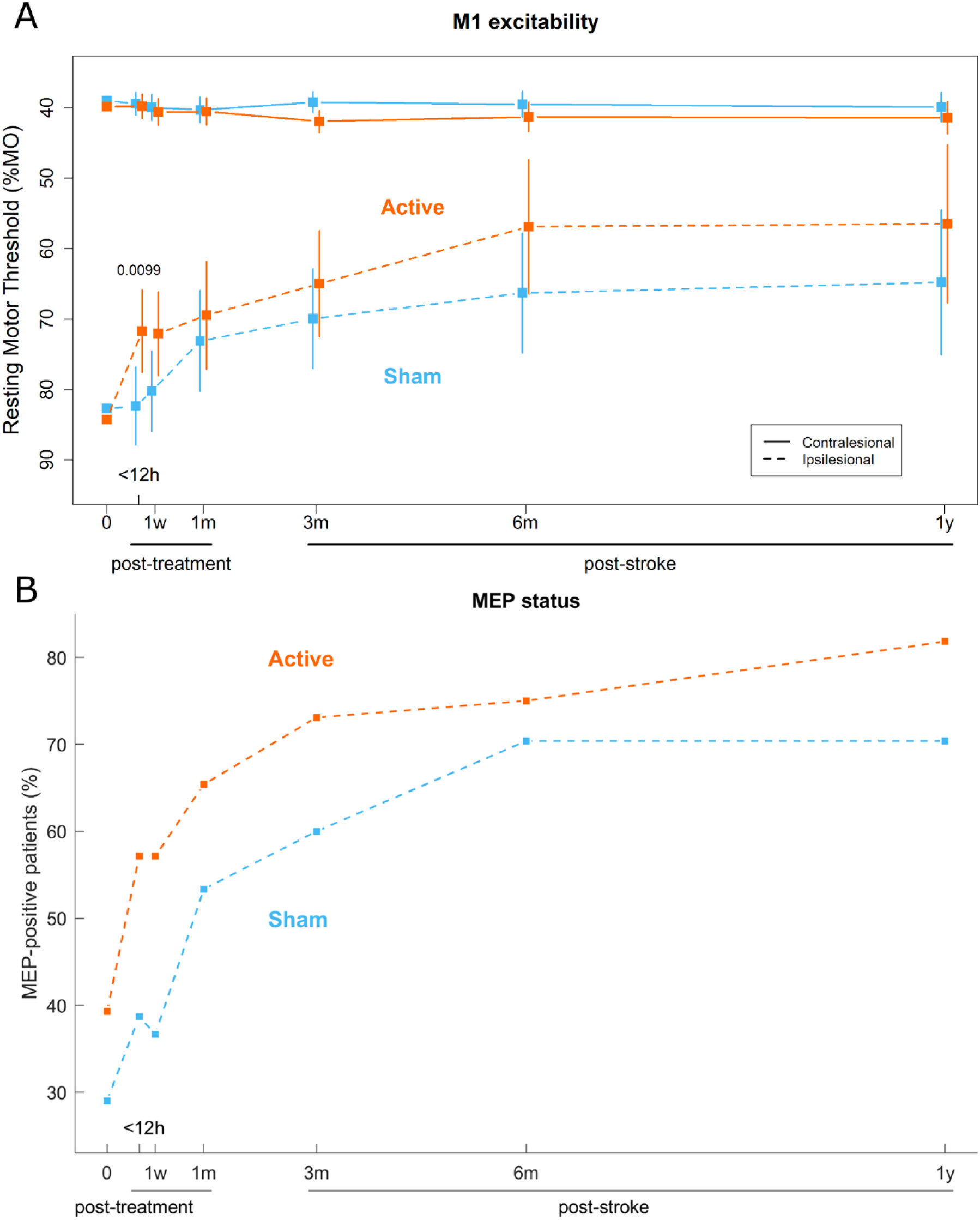
A. Mean and 95% confidence intervals of the ipsi- and contralesional resting motor threshold (RMT) in the active and sham cTBS groups were calculated using a mixed model for repeated measures. Individual data points are shown in supplementary figure S1. The vertical axis is inverted to reflect an increase in M1 excitability along the axis. Figure 3B. The percentage of patients who were MEP-positive in the active and sham cTBS groups. 0: baseline; h: hours; w: weeks; m: months; y: years.

We investigated whether the difference in ipsilesional RMT between the active and sham cTBS group within twelve hours after the series of treatments could be attributed to a change in number of MEP-positive patients. However, there was no difference in the percentage of MEP-positive patients between the active and sham cTBS group within twelve hours after the series of treatments (OR 3.8; 95% CI 0.3 to 53.3, p 0.3193; Figure 3B).

Compared to sham-stimulated patients, patients who received active cTBS had 27% (95% CI −44 to −11; p 0.0030), 25% (95% CI −45 to −5; p 0.0224) and 29% (95% CI −48 to −11; p 0.0038) less overlap with the first MEA within twelve hours post-treatment, at one week post-treatment and at three months post-stroke, respectively (Figure 4A). Ipsilesional MEA size was larger in the active cTBS treatment group compared to the sham stimulation group at six months post-stroke (mean difference 2.52 cm^2^; 95% CI 0.55 to 4.49; p 0.0134; Figure 4B). There were no differences in COG displacement between the active and sham cTBS groups (supplementary figure S4). MEA overlap and change in RMT at the stage of their maximum deviation, i.e. within twelve hours post-treatment were not correlated (r 0.17; p 0.572).

**Figure 4.**
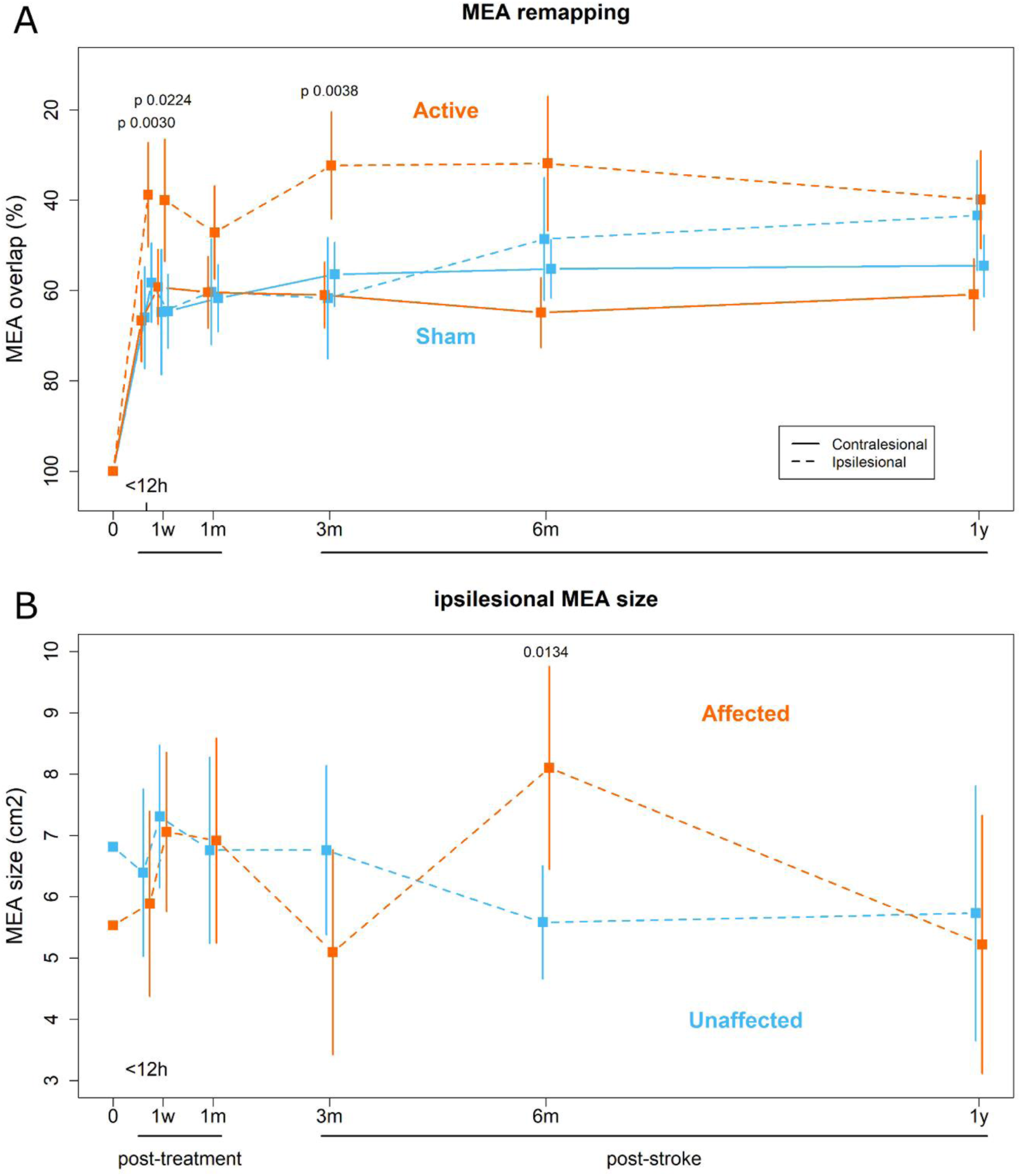
A. Mean and 95% confidence intervals of ipsi- and contralesional motor-eloquent area (MEA) overlap with the first observed MEA in the active and sham cTBS groups calculated using mixed-effects model for repeated measures. Individual data points are shown in supplementary figure S2. The vertical axis is inverted to reflect an increase in MEA remapping along the axis. Figure 4B. Mean and 95% confidence intervals of ipsilesional MEA size in the active and sham cTBS groups calculated using mixed-effects model for repeated measures. Individual data points are shown in supplementary figure S3. 0: baseline; h: hours; w: weeks; m: months; y: years.

Ipsilesional M1 excitability (i.e., RMT) at baseline correlated with outcome on the FMA arm score at baseline (Spearman’s rho 0.66; p < 0.0001; Figure 5A), and ipsilesional M1 excitability within twelve hours post-cTBS correlated with FMA arm score at three months post-stroke (Spearman’s rho 0.59; p < 0.0001; Figure 5C). Ipsilesional M1 excitability at one year post-stroke correlated with outcome on the FMA arm score at one year post-stroke (Spearman’s rho 0.52; p 0.0001; Figure 5E). Ipsilesional M1 excitability did not correlate with the FMA arm score at any of the above timepoints when MEP-negative patients were excluded from analysis (supplementary figure S5).

**Figure 5.**
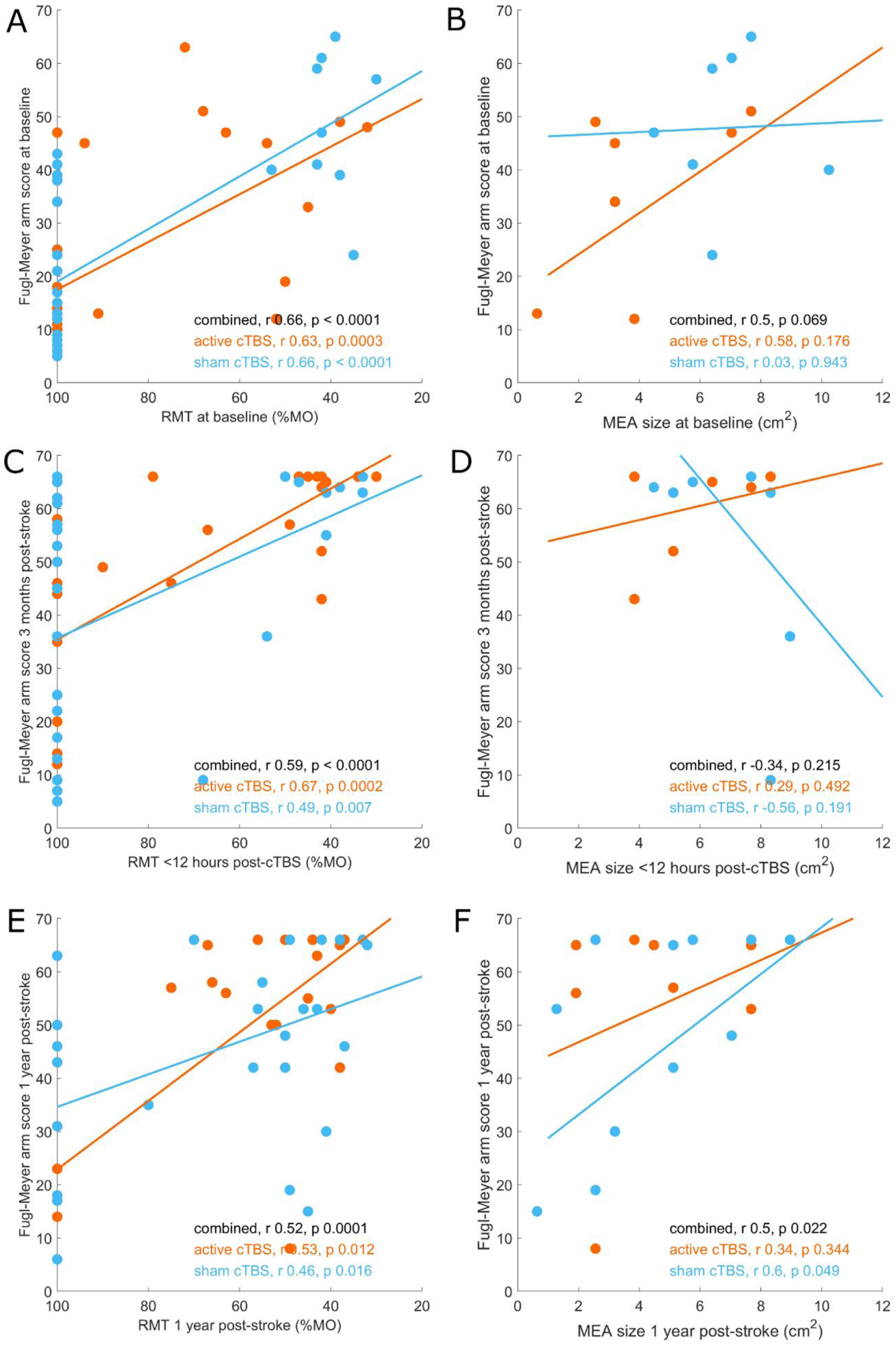
Ipsilesional resting motor threshold (RMT) versus Fugl-Meyer Assessment (FMA) arm score at indicated timepoints, with Spearman’s correlations and an inverted horizontal axis to reflect an increase in M1 excitability along the axis (**A, C, E**). Motor-eloquent area (MEA) size versus FMA arm score at indicated timepoints with Pearson’s correlations (**B, D, E**). Patients in the active cTBS group are shown in orange and patients in the sham cTBS group are shown in blue.

Ipsilesional MEA size at baseline did not correlate with outcome on the FMA arm score at baseline (Pearson’s rho 0.5; p 0.069; Figure 5B), and ipsilesional MEA size within twelve hours post-cTBS did not correlate with FMA arm score at three months post-stroke (Pearson’s rho −0.34; p 0.215; Figure 5D). Ipsilesional MEA size at one year post-stroke correlated with FMA arm score at one year post-stroke (Pearson’s rho 0.5; p 0.022; Figure 5F).

## Discussion

We recently showed in a randomised controlled trial that ten sessions of cTBS of the contralesional M1, combined with upper limb physical therapy, promote upper limb recovery, reduce disability and dependence, and improve quality of life in recovering stroke patients.^15^ Our current study shows normalisation of an interhemispheric balance in M1 excitability, indicated by an increase in ipsilesional M1 excitability with stable contralesional M1 excitability, and a greater extent of ipsilesional MEA remapping, indicated by longitudinally less MEA overlap, in patients who received ten sessions of active cTBS compared to patients who received sham cTBS. These changes primarily took place within the first three months post-stroke, during the presumed window of enhanced plasticity in stroke patients.^26,27^

It has been shown that cTBS can inhibit M1 excitability and increase excitability of the contralateral (non-stimulated) M1.^28^ In stroke patients, this effect, combined with subsequent physical therapy of the affected arm, could potentially facilitate an enduring increase in ipsilesional M1 excitability. We previously found a trend toward a reduction in contralesional M1 excitability during two weeks of cTBS treatment, which was not associated with upper limb function.^15^ We speculate that active cTBS leads to a short-lasting reduction in contralesional M1 excitability, which has largely dissipated the next day. The current study shows that ipsilesional M1 excitability increases within twelve hours after the series of treatments, which cannot be attributed to an increase in the number of MEP-positive patients alone. Ipsilesional M1 excitability post-cTBS was associated with upper limb function at three months post-stroke, which is in line with earlier studies.^29^ We speculate that heightened ipsilesional M1 excitability plays a causal role in upper limb recovery, which may be promoted by cTBS of the contralesional M1.

Rehabilitative training-induced motor recovery, but not spontaneous restoration of motor function, has previously been associated with ipsilesional MEA remapping in non-human primates.^2,30^ We show that remapping can be observed in the ipsilesional MEA in patients (across both treatment arms) who received physical therapy of the upper limb. The observed remapping is specific to the ipsilesional MEA, as the contralesional MEA did not significantly change over time. Additionally, we found that patients who received ten sessions of cTBS, combined with upper limb physical therapy, showed a greater extent of ipsilesional MEA remapping compared to patients who received sham stimulation. This observation is in agreement with previous studies that reported post-stroke remapping of motor representations after treatment.^31,32^

Ipsilesional MEA remapping was characterised by a mean increase in size after treatment. An increase in MEA size associated with improvement in upper limb function has previously been observed after constraint-induced movement therapy.^12,33^ The observed MEA remapping may have occurred through recruitment of normally inactive or latent motor representations or through recruitment of neural circuits that are normally not associated with motor function.^34^ Previous studies have suggested that the MEA may predominantly shift posteriorly after stroke, which could represent recruitment of direct projections from neurons in the postcentral gyrus to spinal or bulbar motor neurons.^35^ However, we observed variable displacement of the ipsilesional MEA in anteroposterior and mediolateral direction after stroke, which is in line with other studies.^27,36^ Therefore, we speculate that the direction of the displacement depends on the location of latent motor representations towards which the MEA could shift, which may differ between stroke patients.

Previous studies reported a relationship between ipsilesional MEA size and upper limb function in the subacute phase^37,38^ and the chronic post-stroke phases.^39^ In the present study, we only identified a relationship between MEA size and upper limb outcome in the chronic phase. We observed a slight decrease in ipsilesional MEA size over the first post-stroke year across both treatment arms. A reduction in MEA of distant upper limb muscle representations has previously been reported in non-human primates that did not receive rehabilitative training.^30^ A possible explanation for this finding in our study could be reduced upper limb use, despite upper limb physical therapy, due to persisting stroke-induced functional limitations in activities of daily living.

The measured increases in ipsilesional M1 excitability and MEA remapping were most prominent within twelve hours after the treatment period. However, these neurophysiological changes were not correlated, suggesting that these are two possible treatment mechanisms that operate independently. However, future studies should elucidate whether enhanced M1 excitability and MEA remapping are a direct cause or rather a consequence of upper limb recovery after stroke.

### Limitations

Our study had limitations. First, TMS-based motor mapping depends on the ability to evoke MEPs upon motor cortex stimulation. Unfortunately, MEPs can typically not be evoked in severely impaired patients in the first weeks to months after stroke,^40^ hampering the collection of data early after stroke in these patients. Therefore, longitudinal changes in MEA characteristics were calculated relative to the first available MEA, potentially reducing statistical power at the early post-stroke time points.

Second, MEP status and ipsilesional excitability in MEP-positive patients were not balanced between the groups at baseline, which may have introduced bias in the statistical analysis. However, the main analysis, including a covariate for baseline ipsilesional excitability, and a sensitivity analysis, including baseline MEP status as an additional covariate, showed that these imbalances were unlikely to affect analysis outcome.

## Conclusion

Ten sessions of cTBS of the contralesional M1 combined with physical therapy lead to heightened ipsilesional M1 excitability and a larger extent of ipsilesional MEA remapping, shortly after the treatment course, reflecting a potential therapeutic mode of action of contralesional cTBS treatment after stroke. Increased ipsilesional M1 excitability and MEA expansion were associated with recovery of the upper limb.

## Data Availability

Deidentified participant data are available from the corresponding author upon reasonable request.

## Acknowledgements

We would like to thank all patients and investigators for their contribution to the B-STARS study.

## Supplemental material

Supplemental material is available.

